# Educational attainment as a modifier of the effect of polygenic scores for cardiovascular risk factors: cross-sectional and prospective analysis of UK Biobank

**DOI:** 10.1101/2021.03.16.21253723

**Authors:** Alice R Carter, Sean Harrison, Dipender Gill, George Davey Smith, Amy E Taylor, Laura D Howe, Neil M Davies

## Abstract

**Background:** Understanding the interplay between educational attainment and genetic predictors of cardiovascular risk may improve our understanding of mechanisms relating educational attainment to cardiovascular disease.

**Methods:** In up to 320 120 UK Biobank participants of White British ancestry (mean age = 57, female 54%), we created polygenic scores for nine cardiovascular risk factors or diseases: alcohol consumption, body mass index, low-density lipoprotein cholesterol, lifetime smoking behaviour, systolic blood pressure, atrial fibrillation, coronary heart disease, type 2 diabetes and stroke. We estimated whether educational attainment modified genetic susceptibility to these risk factors and diseases.

**Results:** On the additive scale, higher educational attainment reduced genetic susceptibility to higher BMI, smoking, atrial fibrillation and type 2 diabetes, but increased genetic susceptibility to higher LDL-C and higher systolic blood pressure.

On the multiplicative scale, there was evidence that higher educational attainment increased genetic susceptibility to atrial fibrillation and coronary heart disease, but no evidence of effect modification was found for all other considered traits.

**Conclusions:** Educational attainment modifies the genetic susceptibility to some cardiovascular risk factors and diseases. The direction of this effect was mixed across traits considered and differences in associations between the effect of the polygenic score across strata of educational attainment was uniformly small. Therefore, any effect modification by education of genetic susceptibility to cardiovascular risk factors or diseases is unlikely to contribute substantially to the mechanisms driving inequalities in cardiovascular risk.

**Key Messages:**

- The role of educational attainment in modifying the effect of polygenic scores for a wide range of cardiovascular risk factors or diseases has not previously been studied
- We explore whether educational attainment modifies the effects of polygenic susceptibility to alcohol consumption, body mass index, low-density lipoprotein cholesterol, lifetime smoking behaviour, systolic blood pressure, atrial fibrillation, coronary heart disease, type 2 diabetes and stroke
- Effect modification by education was observed for some cardiovascular polygenic scores, but not all.
- Effects were not always in the hypothesised direction and were dependent on the scale of analysis.
- Modification of the effect of genetic susceptibility to cardiovascular risk factors or cardiovascular disease by educational attainment is unlikely to contribute substantially to the mechanisms driving inequalities in cardiovascular risk.

## Introduction

Although rates of cardiovascular disease (CVD) have decreased in high income countries, socioeconomically deprived individuals have the greatest risk of disease (1). Most cardiovascular outcomes are complex multifactorial diseases with both environmental and genetic aetiology (2-4). Therefore, it is plausible that socioeconomic position (SEP) may interact with, or modify, genetic susceptibility to CVD.

Large genome-wide association studies (GWAS) have identified many genetic variants that reliably associate with liability to cardiovascular disease and risk factors for CVD (5-7). These can be used to construct polygenic scores, explaining substantial fractions of variation. Tyrrell and colleagues demonstrated that individuals with a higher Townsend deprivation index have an accentuated risk of obesity in genetically susceptible adults (8). Rask-Anderson and colleagues replicated this association, however, they found little evidence that education modified the effect of genetic susceptibility to a high body mass index (BMI) risk on measured BMI (9). Amin and colleagues also found little evidence that education modified genetic susceptibility to BMI in a study using data from the UK and Finland (10).

Whilst educational attainment has been shown to modify the association of cardiovascular risk factors on CVD (1, 11) it is unclear whether educational attainment modifies the effect of genetic susceptibility to a wide range of cardiovascular risk factors. If higher levels of education mitigate some of the genetic risk of cardiovascular risk (‘gene*environment interaction’), this may be one of the mechanisms underlying educational inequalities in cardiovascular disease (12).

## Methods

### UK Biobank

The UK Biobank recruited 503 317 adults from around the UK between 2006 and 2010, aged 37 to 73 (13). Participants attended baseline assessment centres involving questionnaires, interviews, anthropometric, physical and genetic measurements (13, 14). In this analysis, we use up to 320 120 individuals of White British ancestry (sFigure 1).

### Educational attainment

At baseline, participants reported highest qualification achieved, which was converted to the International Standard Classification for Education (ISCED) coding of educational attainment (sTable 1) (15).

### Cardiovascular risk factors and cardiovascular disease

Cardiovascular risk factors were included in our study if there is evidence from randomized controlled trials, Mendelian randomisation studies, or clinical studies, that they are a causal risk factor for CVD (see sTable 2 for studies) and have suitable GWAS summary statistics available. Additionally, we included polygenic scores for a number of CVD subtypes. In total, 9 polygenic scores were included in our analyses, these were; alcohol consumption (16), BMI (17), type 2 diabetes (18), low density lipoprotein cholesterol (LDL-C) (19), lifetime smoking behaviour (20) and systolic blood pressure (21). Polygenic scores for cardiovascular subtypes included in analyses were atrial fibrillation (5), coronary heart disease (CHD) (6) and stroke (7).

Cardiovascular risk factors were measured at baseline assessment centres, whilst incident cardiovascular outcomes (atrial fibrillation, CHD, stroke and type 2 diabetes) were determined by linked mortality records and hospital inpatient records (see sTable 3). A full description of how each risk factor/outcome was measured, both phenotypically and genetically, is presented in the Supplementary Methods.

### Deriving polygenic scores

Summary statistics of the associations of the single-nucleotide polymorphisms (SNPs) with each cardiovascular risk factor/outcome were downloaded from MR-Base (22) or the relevant GWAS. We used the most recent GWAS for each risk factor/ outcome excluding UK Biobank participants to avoid bias by sample overlap. The 1000 genomes project was used to find proxy SNPs in LD with SNPs not found in UK Biobank. Pruning of SNPs was carried out using the clump command in PLINK using an r^2^ parameter of 0.25 and a physical distance threshold for clumping of 500kB. Polygenic scores were constructed using a range of p-value thresholds p≤5×10^−8^ (genome-wide significant), ≤0.05, and ≤0.5. As the p-value threshold increases, the variance explained by the polygenic score typically increases. However, increasing the numbers of SNPs increases the risk of pleiotropy and false positive effects. The pruned SNPs from each GWAS were harmonised with the SNPs from UK Biobank, aligning the effect estimates and alleles. Any SNPs that could not be harmonised, palindromic SNPs (where alleles on the forward and reverse strand are read the same) or triallelic SNPs were excluded from polygenic scores. The polygenic scores were created by multiplying the number of effect alleles for each participant by the association of the SNP with the phenotype in each GWAS, then summed across all SNPs associated with each phenotype. All polygenic scores were standardized for use in analyses and reflect a one standard deviation (SD) change.

Main analyses are presented using polygenic scores derived at the genome-wide significance threshold with other polygenic score thresholds presented in the supplement.

### Exclusion criteria

Reverse causality can introduce bias when the temporality of the exposure and outcome is mis-specified and the outcome itself affects the exposure (23). Although a cardiovascular diagnosis in later life can not alter genetic variants defined at conception, and indeed is unlikely to change educational attainment typically determined by early adulthood, a diagnosis may lead to behavioural or lifestyle changes which change the relative importance of the polygenic score in determining the outcome state. To avoid bias by reverse causality, analyses of cardiovascular outcomes were carried out prospectively and individuals were excluded if they had experienced at least one diagnosis of any of the outcomes considered in analyses at, or before, baseline (atrial fibrillation, CHD, stroke and type 2 diabetes). Additionally, participants were excluded if they had experienced any one of myocardial infarction, angina, transient ischaemic attack, peripheral arterial disease, familial hypercholesterolaemia, type 1 diabetes and chronic kidney disease, which can all result in statins being prescribed to prevent CVD which may lead to behaviour change and therefore reverse causality (24). All diagnoses were ascertained through linkage to mortality data and hospital inpatient records, with cases defined according to ICD-9 and ICD-10 codes (sTable 4).

Quality control of the genetic data was carried out according to the Medical Research Council Integrative Epidemiology Unit quality control pipeline, described in full previously (25). In brief, individuals were excluded if their genetic sex differed to their gender reported at baseline or for having aneuploidy of their sex chromosomes (non-XX or -XY chromosomes). Further individuals were excluded for extreme heterozygosity or a substantial proportion of missing genetic data. Related individuals were excluded removing those related to the greatest number of other participants, until no related pairs were left (25). This exclusion list was derived in-house using an algorithm applied to the list of all the related pairs provided by UK Biobank (3^rd^ degree or closer) (sFigure 1). In addition, individuals were excluded if they had withdrawn from UK Biobank or were, or may be, pregnant at baseline.

Individuals were excluded if they were missing data for education, age and sex. Individuals were excluded from specific analyses if they were missing phenotypic measurements of the risk factor/outcome under consideration (see sFigure 1).

## Statistical Analysis

### Association of educational attainment with outcomes

Multivariable linear regression (adjusting for age and sex) was carried out to estimate the association between educational attainment and cardiovascular risk factors/diseases.

### Association between each polygenic score and observed phenotype

For each of the cardiovascular risk factors/diseases, we estimated the association between each polygenic score and the corresponding phenotypic using multivariable linear regression, adjusting for age, sex and 40 genetic principal components to control for population structure. For continuous cardiovascular risk factors, measures were standardised, so estimates reflect the mean difference in SD of the phenotype for a one SD higher polygenic score. For binary outcomes, estimates reflect the risk difference or odds ratio of the outcome per one SD higher polygenic score.

### Effect modification by educational attainment on polygenic scores for cardiovascular risk

To test for effect modification, the linear model was stratified by years of educational attainment. To estimate the magnitude and direction of the effect modification, an interaction term was included in the linear model (e.g., polygenic score*education [continuous]). Analyses were adjusted for age, sex and 40 genetic principal components. As effect modification is scale dependent and, by definition, with two causal risk factors an interaction will be present on at least one of the additive and multiplicative scales, tests of effect modification were carried out on both the additive and multiplicative scale (26).

## Secondary Analyses

All analyses were replicated for polygenic scores at P value thresholds of ≤0.05 and ≤0.5.

## Data and code availability

The data used in this study has been archived with the UK Biobank study and was carried out under approved project 10953. The analysis code used is available at github.com/alicerosecarter/gxe_cv_riskfactors.

## Results

### UK Biobank cohort

Eligible UK Biobank participants (55% female) had a mean age of 57 (standard deviation [SD] = 8.00 years). A higher proportion of participants (33%) left school after 20 years (equivalent to obtaining a degree), compared with those who left school after 7 years (equivalent to no formal qualifications) (16%) (Table 1).

**Table 1:** Descriptive characteristics of the main analysis sample compared with all individuals in UK Biobank at baseline

| Variable | Analysis sample |  |  | Full UK biobank* |  |
| --- | --- | --- | --- | --- | --- |
|  | (N = 320 120) |  |  | (N = 502 156) |  |
| Continuous variables | N |  | Mean (SD) | N | Mean (SD) |
| Age | 320 120 |  | 56.66 (8.00) | 502 156 | 56.54 (8.09) |
| Drinks per week | 318 300 |  | 8.17 (9.05) | 497 917 | 7.79 (9.05) |
| BMI | 319 201 |  | 27.3 (4.72) | 499 065 | 27.43 (4.8) |
| LDL-C | 304 700 |  | 3.61 (0.86) | 468 390 | 3.56 (0.87) |
| Systolic blood pressure | 292 277 |  | 138.16<br>(18.58) | 456 647 | 137.79<br>(18.62) |
| Smoking (lifetime behaviour) | 301 684 |  | 0.32 (0.66) | 318 112 | 0.34 (0.67) |
| Categorical variables | N |  | Frequency (%) | N | Frequency (%) |
| Sex | Female | 320 120 | 175 108 (55) | 502 156 | 273 025 (54) |
| Years of education | 7 years | 320 120 | 52012 (16) | 493 033 | 84648 (17) |
|  | 10 years |  | 54899 (17) |  | 82357 (17) |
|  | 13 years |  | 17355 (5) |  | 26857 (5) |
|  | 15 years |  | 39144 (12) |  | 58271 (12) |
|  | 19 years |  | 51418 (16) |  | 77668 (16) |
|  | 20 years |  | 105292 (33) |  | 163232 (33) |
| Atrial fibrillation (incident) | Control | 316 912 | 307352 (97) | 495 772 | 480007 (97) |
|  | Case |  | 9560 (3) |  | 15765 (3) |
| Coronary artery disease (incident) | Control | 317 055 | 302574 (95) | 481 533 | 458689 (95) |
|  | Case |  | 14481 (5) |  | 22844 (5) |
| Type 2 diabetes (incident) | Control | 316 406 | 305327 (96) | 492 726 | 472098 (96) |
|  | Case |  | 11079 (4) |  | 20628 (4) |
| Stroke (incident) | Control | 320 120 | 314191 (98) | 497 151 | 487084 (98) |
|  | Case |  | 5929 (2) |  | 10067 (2) |

For a P value of ≤5×10^−8^, the polygenic scores explained between 0.06% (atrial fibrillation) and 14% (systolic blood pressure) of variance in the phenotypes (sTable 5).

### Association between educational attainment, polygenic scores and cardiovascular risk factors use

Educational attainment was associated with all cardiovascular risk factors/diseases, except for LDL-C, although for all outcomes the effect was small (sTable 6). Except for alcohol consumption, higher educational attainment led to a reduction in the mean difference of all risk factors/diseases (sTable 6).

### Effect modification by educational attainment of genetic susceptibility to cardiovascular risk factors

For most polygenic scores, there was evidence that educational attainment modified the effect of the polygenic score on either the additive or multiplicative scale (Figures 1-3 and sTables 7 and 8). The exception was alcohol consumption, for which there was little evidence on either scale.

**Figure 1:**
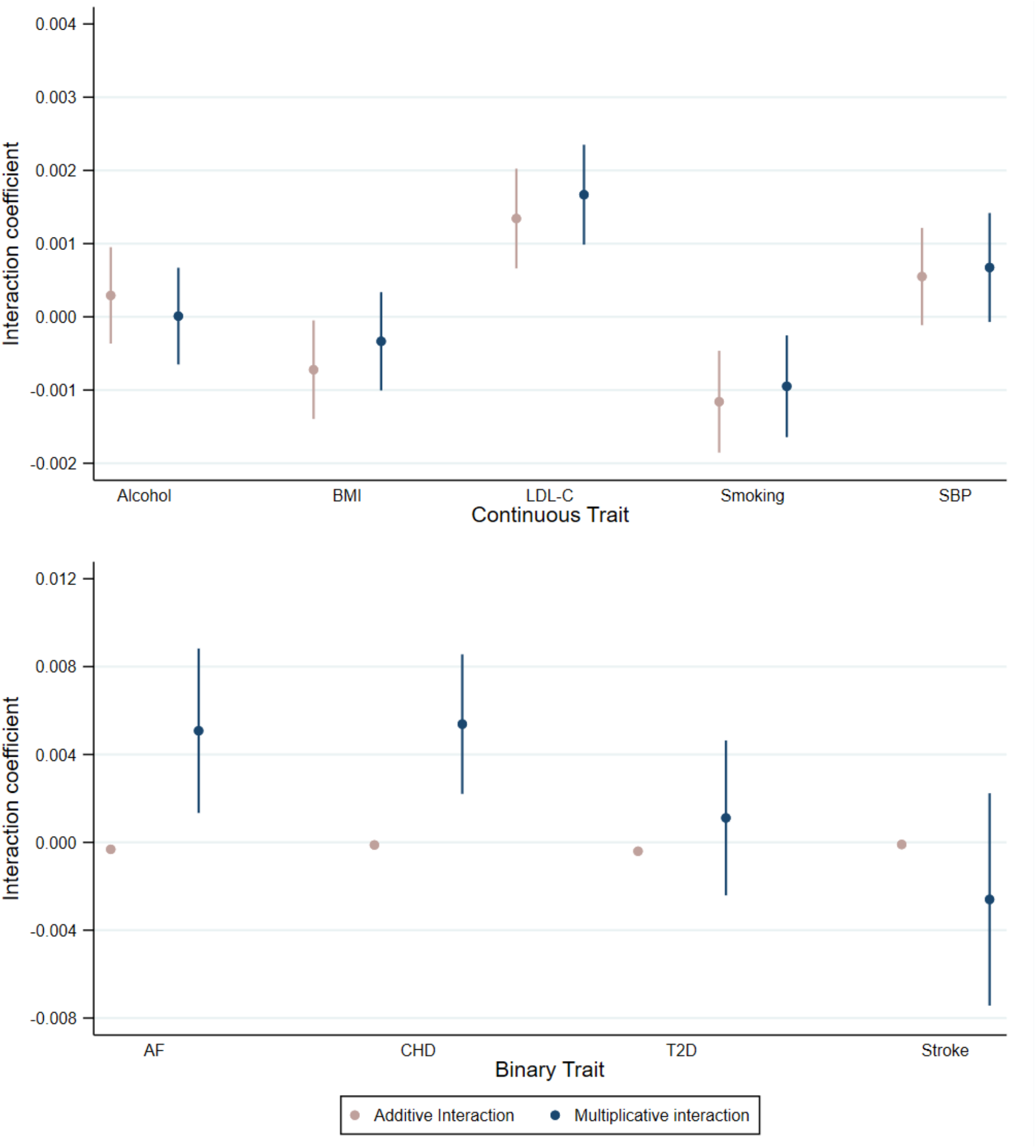
Coefficient for educational attainment as an effect modifier of polygenic susceptibility to cardiovascular risk factors or diseases on the additive and multiplicative scale Analyses adjusted for age, sex and 40 genetic principal components Alcohol = drinks per weekly BMI = body mass index; LDL-C = Low density lipoprotein cholesterol; smoking = lifetime smoking behaviour; SBP = systolic blood pressure; AF = Atrial fibrillation; CHD = Coronary heart disease; T2D = Type 2 diabetes Analyses for binary outcomes on the multiplicative scale are presented as log odds ratios

**Figure 3:**
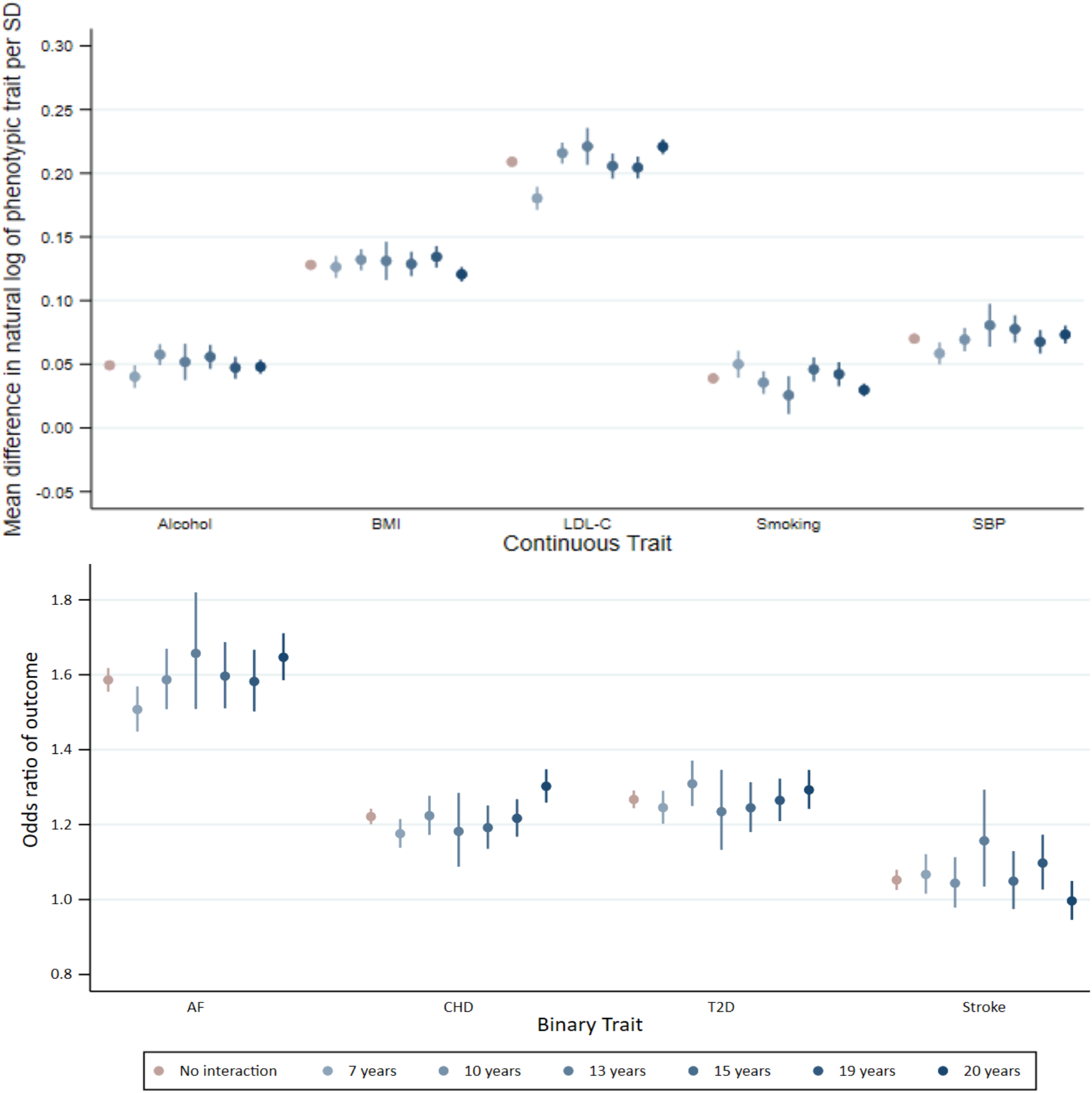
Association between polygenic scores for susceptibility to cardiovascular risk and phenotypic measure of each risk factor, stratified by educational attainment demonstrating effect modification on the multiplicative scale Analyses adjusted for age, sex and 40 genetic principal components Alcohol (drinks per week) P_EM_ = 0.976; body mass index (BMI) P_EM_ = 0.330; low-density lipoprotein cholesterol (LDL-C) P_EM_ = 1.63×10^−6^; lifetime smoking behaviour P_EM_ = 0.008; systolic blood pressure (SBP) P_EM_ = 0.076 Atrial fibrillation (AF) P_EM_ = 0.008; coronary heart disease (CHD) P_EM_ = 8.94×10^−4^; type 2 diabetes (T2D) P_EM_ = 0.537; stroke P_EM_ = 0.292 EM = effect modification

On the additive scale, higher educational attainment protected against genetic susceptibility to higher BMI, smoking, atrial fibrillation and type 2 diabetes (Figure 1 and Figure 2). For example, a one SD increase in polygenic score for smoking increased mean difference in lifetime smoking by 0.05 SD (95% CI: 0.04 to 0.06) for those with 7 years education and by 0.03 SD (95% CI: 0.02 to 0.03) for people with 20 years of education (Figures 1 and 2 and sTable 7) (P_effect modification_ = 0.001).

**Figure 2:**
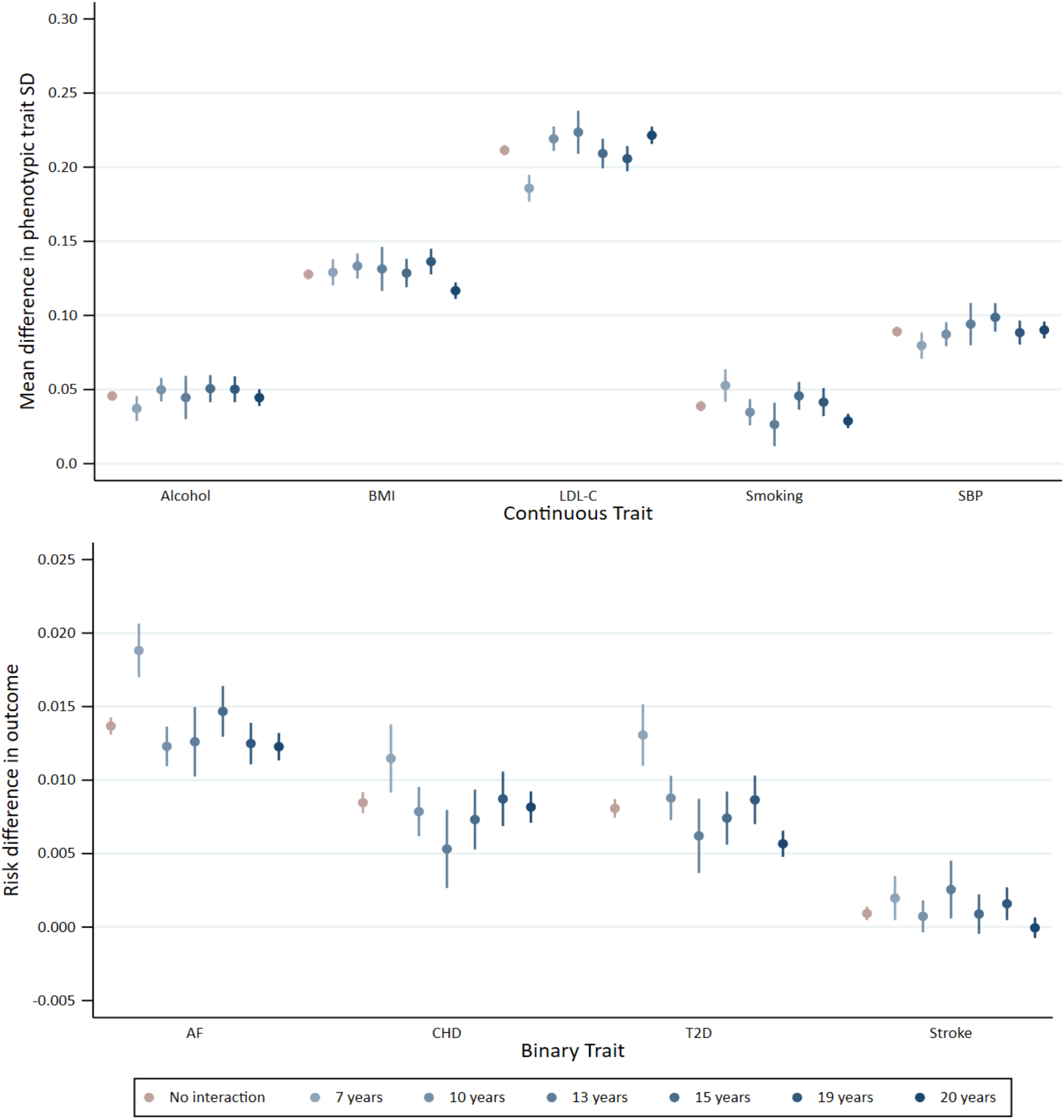
Association between polygenic scores for susceptibility to cardiovascular risk and phenotypic measure of each risk factor, stratified by educational attainment demonstrating effect modification on the additive scale Analyses adjusted for age, sex and 40 genetic principal components Alcohol (drinks per week) P_EM_ = 0.384; body mass index (BMI) P_EM_ = 0.036; low-density lipoprotein cholesterol (LDL-C) P_EM_ = 1.12×10^−4^; lifetime smoking behaviour P_EM_ = 0.001; systolic blood pressure (SBP) P_EM_ = 0.104 Atrial fibrillation (AF) P_EM_ = 9.03×10^−8^; coronary heart disease (CHD) P_EM_ = 0.103; type 2 diabetes (T2D) P_EM_ = 3.23×10^−10^; stroke P_EM_ = 0.036

Also on the additive scale, higher educational attainment increased genetic susceptibility to LDL-C and systolic blood pressure. For example, for those with 7 years of education an increase of one SD in the polygenic score for LDL-C increased mean LDL-C by 0.19 SD (95% CI: 0.18 to 0.19), compared to 0.22 SD (95% CI: 0.22 to 0.23) for people with 20 years of education(P_effect modification_ = 1.12×10^−4^) per SD increase in polygenic score (Figures 1 and 2 and sTable 7).

On the multiplicative scale, there was evidence that higher educational attainment increased genetic susceptibility to atrial fibrillation and CHD. For example, for a one SD increase in atrial fibrillation polygenic score, the odds ratio for atrial fibrillation in individuals with 7 years of education was 1.59 (95% CI: 1.45 to 1.57) and for people with 20 years of educational attainment the odds ratio was 1.65 (95% CI: 1.59 to 1.71) (P_effect modification_ = 9.03×10^−8^) (Figures 1 and 3 and sTable 8). There was little evidence of modification by education on the multiplicative scale for all other polygenic scores.

For all outcomes, the size of the coefficients for effect modification was small. Non-linear effects by strata of educational attainment were observed for a number of outcomes (Figures 2 and 3).

### Secondary analyses

Analyses using more liberal P-value thresholds for the polygenic score were broadly consistent with the main genome-wide results. Similar directions of effect were observed, for example on the additive scale a one unit increase in educational attainment protected against genetic susceptibility to BMI and lifetime smoking behaviour (sTable 9 and sTable 10).

## Discussion

In this analysis of UK Biobank participants, we found evidence that educational attainment modified the risk of genetic susceptibility to some, but not all, cardiovascular risk factors/diseases. We had hypothesised that higher levels of education would mitigate genetic susceptibility to cardiovascular risk, but in several cases the effect modification was in the other direction, i.e., higher education accentuated genetic predisposition. Furthermore, the magnitude of the differences in associations between polygenic scores and cardiovascular risk factors/disease across levels of educational attainment was small in all cases, and the evidence for effect modification was often scale dependent. These results suggest that modification of the effect of polygenic scores by educational attainment is unlikely to play a substantial role in the generation of educational inequalities in cardiovascular disease.

## Results in context

A number of studies have sought to identify the interplay between genetic susceptibility to cardiovascular risk factors with a range of lifestyle and environmental factors (27-32). However, few have considered the role of SEP interacting with genetic risk.

Two recent studies using UK Biobank have demonstrated that a greater Townsend deprivation index accentuated the genetic risk of obesity (8, 9). However, the previous literature has not found evidence that education modifies the genetic risk of obesity (9, 10). We have expanded on this here by exploring the extent to which education modifies polygenic susceptibility to a wide range of cardiovascular risk factors, rather than focussing on one risk factor. In contrast to the previous literature, we found evidence that educational attainment modifies genetic susceptibility to BMI. This may be related to power, where previous studies have used smaller sample sizes to estimate interactions.

A further explanation for these differences could be because of the education definition used. Here, we converted highest educational qualification to ISCED years of schooling, however previous research has defined education as age of completing full time education (9) and highest qualification (10).

## Strengths and weaknesses, and caveats to the analysis of effect modification

There are a number of strengths in this study. Much of the previous literature on gene*environment interactions in CVD rely on candidate gene style studies (31, 33, 34), which have often been shown to be spurious (35). Here, we have used polygenic scores for nine cardiovascular risk factors/diseases. Whilst candidate gene studies typically focus on a single genetic variant, or small group of (common) genetic variants that individually explain a large(r) amount of the variance in the trait, polygenic scores include a large number of genetic variants which each explain a small amount of the variation, but cumulatively explain a large amount (36, 37). For most diseases, including CVD, polygenic inheritance of these common variants plays a greater role than rare monogenic mutations (37, 38). Therefore, the broad measure of genetic susceptibility used here is likely to represent a greater number of biological pathways for the aetiology of cardiovascular disease.

Additionally, we created polygenic scores at a range of P value thresholds. At a more stringent threshold (e.g., P≤5×10^−8^) the genetic variants included are less likely to be pleiotropic (i.e., also associated with different phenotypes), but the variance explained by the polygenic score may be lower than with a more liberal threshold (e.g., P≤0.5). Additionally, less stringent clumping thresholds were used to improve polygenic prediction, but this may introduce pleiotropic SNPs.

The lack of evidence for effect modification between education and the polygenic score for alcohol consumption observed in this study could be due to insufficient power to detect an interaction or because of the way the variable was defined. For example, alcohol consumption was defined as drinks per week, but type of alcoholic drink consumed may be an important factor which was not accounted for. Additionally, alcohol consumption was self-reported by participants, which is prone to recall bias (39, 40). If this recall bias is differential by strata of educational attainment, this may lead to a masking of any effect modification between educational attainment and genetic susceptibility to alcohol consumption on actual alcohol consumed. Alternatively, different patterns of drinking may occur in different strata of educational attainment. For example, it has previously been shown that individuals of lower SEP are more likely to drink to extreme levels (41), but individuals of higher SEP consume similar or even greater amounts of alcohol (42).

Low statistical power reduces the chance of detecting a true effect should one exist (43). Although some power calculators have been developed to calculate power in gene*environment interaction analyses (44), to our knowledge none have been developed for use with polygenic scores. Therefore, we cannot calculate the theoretical power of these analyses. However, it is possible that we did not detect effect modification by education on the effects of alcohol consumption due to insufficient power to detect an interaction.

Studies of effect modification can be biased by reverse causality and confounding. Where possible, for example with genetic susceptibility to cardiovascular diagnoses, we restricted analyses to incident cases to avoid bias from behaviour changes following previous disease events. Genetic variants are determined at conception, and therefore not affected by unmeasured later life confounding factors. However, they can be confounded by population structure (45). In this analysis, we controlled for genetic principal components to minimise bias due to this.

One limitation is the generalisability of these results to other populations. UK Biobank is not representative of the wider UK population (13). UK Biobank participants are typically more highly educated and of a higher SEP. Therefore, evidence of that education modifies the effect of polygenic scores in this sample may be due to collider bias caused by non-random selection into the study (46).

Although we have identified evidence to support that education modifies the effect of polygenic scores for some cardiovascular risk factors, these effects may differ for alternative measures of SEP. Likewise, should different definitions of the outcome variables be used, e.g., smoking initiation as opposed to lifetime smoking behaviour, the observed evidence of effect modification may change.

These results do not specifically identify what it is about educational attainment that modifies genetic susceptibility to cardiovascular risk factors/outcomes. For example, remaining in education may lead to an increased knowledge of the smoking harms, even if they have genetic variants increasing their susceptibility to heavier smoking (47).

Where educational attainment increased genetic susceptibility to cardiovascular disease diagnoses, such as atrial fibrillation, it is possible these differences are observed due to differences in rates of diagnosis, which may independently contribute to cardiovascular inequalities. Whilst risk factors such as BMI and smoking were measured near universally in participants at baseline, cardiovascular diagnoses were ascertained through linkage to hospital inpatient records. This may therefore reflect differential diagnosis across strata of educational attainment.

## Interpreting analyses of interaction and effect modification

The terms interaction and effect modification are often used interchangeably in modern epidemiology. Whilst statistically the same, the distinction can be made where an interaction is defined in terms of the effects of two causal risk factors, whereas effect modification specifies that the effect of one causal risk factor varies by strata of a second factor, the effect of which on the outcome is not necessarily causal (48). We have used the term effect modification throughout this analysis, where we specifically hypothesise that the effect of the polygenic scores vary by strata of educational attainment.

Interaction and effect modification have often been dichotomized into “biologic interaction” and “statistical interaction” (49, 50). Biologic interaction is said to be a deviation from an additive effect of two causal risk factors on the risk difference of the outcome. However, the term biologic interaction has been criticised for being difficult to interpret and giving potentially misleading assurances about causal biological mechanisms that have not been assessed (50).

Statistical interaction is described as the deviation from the expected effect of two joint risk factors, under the assumption the risk factors are independent, either on the additive or the multiplicative scale (50). When two risk factors are causal, as is the case in our analyses, there should always be evidence of an interaction on at least one of the scales, therefore we present results on both the additive and the multiplicative scales (26). This is an important distinction from previous analyses, which have typically only reported results on the additive scale (8, 9).

Importantly for public health relevance is to interpret the magnitude of any differences in associations between the effect of the polygenic scores across strata of education. These were uniformly small in this analysis and the direction of the effect was not consistent. This indicates that any effect modification by educational attainment on the effect of genetic susceptibility to cardiovascular risk factors/disease is unlikely to contribute to the mechanisms driving inequalities in cardiovascular risk.

## Conclusions

In this study we have found that educational attainment modifies genetic susceptibility to a number of cardiovascular risk factors. The direction of this effect was mixed, and the size of the effect modification coefficients were small. This suggests that effect modification by educational attainment on the effect of genetic susceptibility to cardiovascular risk factors or cardiovascular disease is unlikely to contribute to the mechanisms driving inequalities in cardiovascular risk.

## Acknowledgements

This publication is the work of the authors, who serve as the guarantors for the contents of this paper. This work was carried out using the computational facilities of the Advanced Computing Research Centre - http://www.bris.ac.uk/acrc/ and the Research Data Storage Facility of the University of Bristol - http://www.bris.ac.uk/acrc/storage/. This research was conducted using the UK Biobank Resource using application 10953.

Quality Control filtering of the UK Biobank data was conducted by R.Mitchell, G.Hemani, T.Dudding, L.Corbin, S.Harrison, L.Paternoster as described in the published protocol (doi: 10.5523/bris.1ovaau5sxunp2cv8rcy88688v). The MRC IEU UK Biobank GWAS pipeline was developed by B.Elsworth, R.Mitchell, C.Raistrick, L.Paternoster, G.Hemani, T.Gaunt (doi: 10.5523/bris.pnoat8cxo0u52p6ynfaekeigi)

