## Supplementary material for "Educational attainment as a modifier of the effect of polygenic scores for cardiovascular risk factors: cross-sectional and prospective analysis of UK Biobank"

### Supplementary methods

##### Alcohol consumption

Alcohol consumption was defined as the number of drinks consumed per week. At baseline assessment centres, participants were asked to described current drinking status (current, former or never) and estimate their current alcohol intake. Of those reporting a current frequency of at least once or twice a week, they were asked to estimate their current average weekly intake of different alcohol beverages. These were summed together to estimate an average number of drinks per week. Individuals reporting a current intake of “one to three times a month” or less frequently, were assumed to have a weekly intake of 0. This variable has been described in detail previously (1).

Summary statistics from the GWAS and Sequencing Consortium of Alcohol and Nicotine use (GSCAN) GWAS of drinks per week were used for the polygenic score (2). This GWAS included predominantly European participants, excluding participants from UK Biobank to avoid sample overlap.

##### BMI

Baseline measures of height and weight were used to calculate BMI (kg/m$^2$).

Summary statistics for use in the polygenic score for BMI came from the Genetic Investigation of Anthropometric Traits (GIANT) Consortium genome-wide association study (GWAS) analysis of 339 224 individuals with European ancestry (3). This is the most recent GWAS of BMI not including UK Biobank, where overlapping samples for the discovery and analysis dataset can lead to inflated effect estimates.

##### Low density lipoprotein cholesterol

Non-fasting measures of low-density lipoprotein cholesterol were measured using enzymatic assays (Backman Coulter AU5800). UK Biobank corrected serum data for laboratory dilution effects and were excluded if they did not pass UK Biobank quality control (4).

Summary statistics for use in the polygenic score came from the Global Lipids Genetics consortium, which included 188 577 males and females of predominantly European ancestry (5).

##### Smoking

A measure of lifetime smoking was constructed in the UK Biobank from self-reported age at initiation, age at cessation and cigarettes per day. From this information, smoking duration and time since cessation were calculated. The lifetime smoking measure further includes a simulated constant (half-life) which captures the exponentially decreasing effect of cigarettes on health over time. Aspects of smoking behaviour were combined into one score ranging from 0 (for non-smokers) to 4.00 (mean = 0.33, standard deviation = 0.67). Full details of score construction can be found elsewhere (6). The main advantage of using this measure of smoking is that it is a continuous measure, improving statistical power, and it considers all aspects of smoking which may affect health, e.g. duration of smoking and smoking heaviness.

We carried out a split sample GWAS of lifetime smoking in UK Biobank to identify genetic variants associated with lifetime smoking to use in a polygenic score. We included 318 147 participants with White British ancestry, who were randomly assigned to one of two samples. In each half of the eligible participants, the GWAS was conducted, which was used to derive the polygenic score in the opposing sample so as to avoid sample overlap which can inflate genetic estimates. This split sample GWAS of lifetime smoking has previously been used in a polygenic score and described in detail (7). The estimates from each sample were meta-analysed to create a single estimate of interaction for smoking.

##### Systolic blood pressure

The mean from two resting automated measures of systolic blood pressure, measured using an Omron HEM-7105IT digital blood pressure monitor at baseline assessment centres was used for phenotypic measurements.

We carried out a split sample GWAS of systolic blood pressure in UK Biobank to identify genetic variants for use in the polygenic score. For individuals who reported taking antihypertensive medication to UK Biobank study nurses, we added 10mm Hg to the phenotypic measurement of systolic blood pressure (8). This GWAS was conducted as described previously for smoking and has been described in detail previously (7). The estimates from each sample were meta-analysed to create a single estimate of interaction for systolic blood pressure.

##### Atrial Fibrillation

Atrial fibrillation events were ascertained through linkage to mortality data and hospital episode statistics (HES) and Scottish morbidity records (SMR), referred to jointly as hospital inpatient records. Cases were defined according to ICD-9 and ICD-10 codes (see sTable 2 for ICD codes used in case definition). Date of diagnoses are provided by HES data, which was linked with the date of assessment centre provided by UK Biobank to identify incident and prevalent cases.

Summary statistics for use in the polygenic score were from a 2012 GWAS of 59 133 individuals (6 707 cases) of European ancestry (9).

##### Coronary heart disease

Coronary heart disease (CHD) events were ascertained through linkage to mortality data and hospital inpatient records, with cases defined according to ICD-9 and ICD-10 codes (see sTable 2 for ICD codes used in case definition) (10). Date of diagnoses are provided by hospital inpatient records, which was linked with the date of assessment centre provided by UK Biobank to identify incident and prevalent cases.

Summary statistics from the most recent GWAS for CHD not including UK Biobank were used for deriving the polygenic score (11). A total of 184 305 individuals (60 801 cases) were included in this GWAS of predominantly European descent.

##### Diabetes

Type 2 diabetes was ascertained by linkage to HES data (sTable 2), with date of diagnosis defined by hospital inpatient records. Additionally, individuals were defined as diabetes if they had reported to UK Biobank study nurses that they had ever had diabetes diagnosed by a doctor (variable 2443). This variable does not distinguish between type 1 and type 2 diabetes, however individuals with a hospital inpatient record for type 1 diabetes were excluded from analyses and in this adult population new diagnoses are more likely to be type 2 diabetes. Individuals were defined as a prevalent case if they reported a diagnosis at baseline assessment centres (variable n_2443_0_0). Incident cases were defined as those who reported a diagnosis at follow up clinics (variable n_2443_1_0 and variable n_2443_2_0), with no previous diagnosis reported (although only a subset of individuals have follow up measures).

Summary statistics of 158 808 European individuals (26 276 Cases) from the DIAbetes Genetics Replication And Meta-analysis (DIAGRAM) Consortium GWAS of type 2 diabetes were used for the polygenic score (12).

##### Stroke

Stroke events (all subtypes) were ascertained through linkage to mortality data and HES data, with cases defined according to ICD-9 and ICD-10 codes (supplementary table 2) (10). Date of diagnoses are provided by hospital inpatient records, which was linked with the date of assessment centre provided by UK Biobank to identify incident and prevalent cases.

For the polygenic score, summary statistics for all subtypes of stroke were obtained from the MEGASTROKE consortium, consisting of 521 612 males and females (67 162 cases) of predominantly European ancestry (13).

### Supplementary figures

sFigure 1: Study flow chart

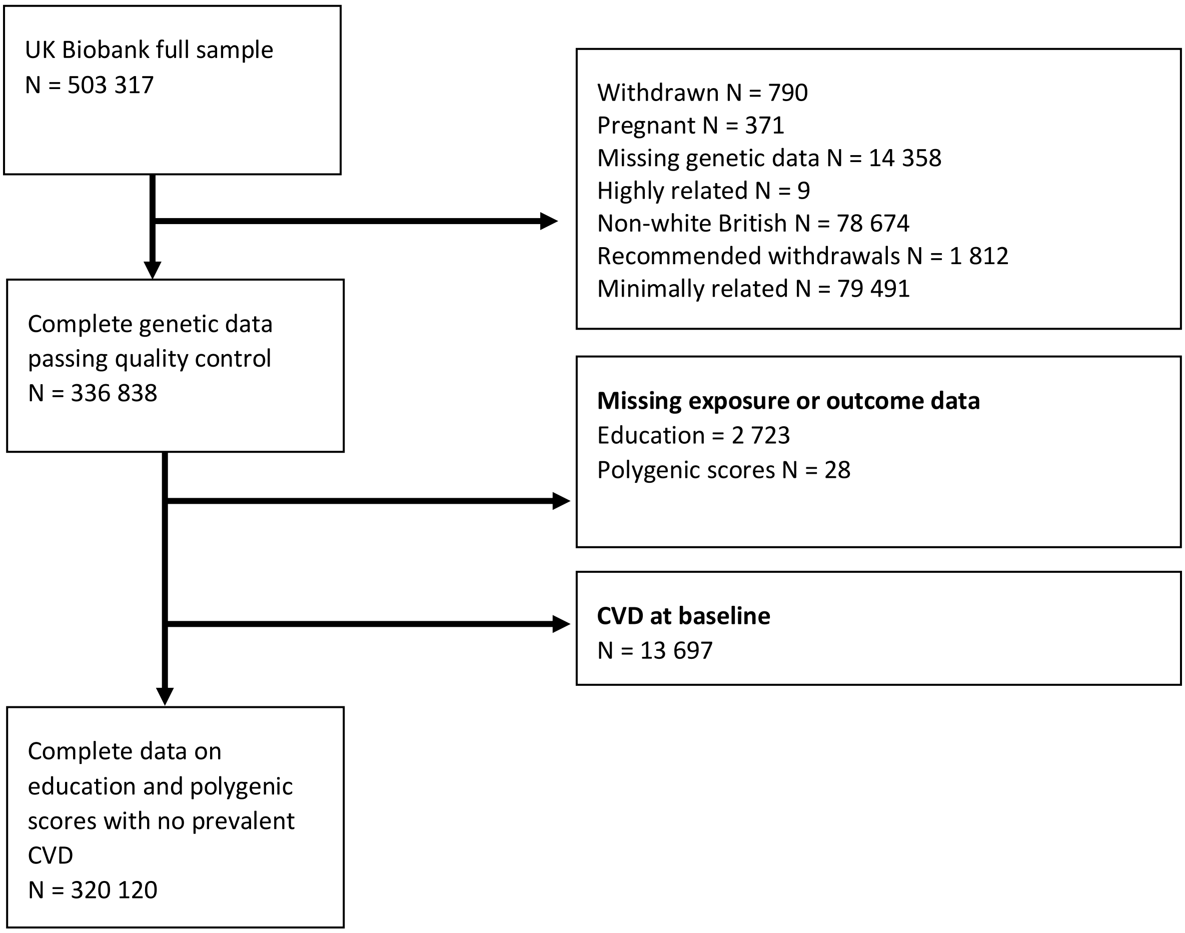

### Supplementary tables

sTable 1: International Standard Classification for Education (ISCED) definitions

| Qualification (As reported in UK Biobank) | ISCED | Years of education |
|---|---|---|
| College or University degree | 5 | 20 |
| NVQ or HND or HNC or equivalent | 5 | 19 |
| Other prof. qual. eg: nursing, teaching | 4 | 15 |
| A levels/AS levels or equivalent | 3 | 13 |
| O levels/GCSEs or equivalent | 2 | 10 |
| CSEs or equivalent | 2 | 10 |
| None of the above | 1 | 7 |
| Prefer not to answer | Excluded | |

sTable 2: Studies providing causal evidence for cardiovascular risk factors included

| Risk factor | Study | Study design |
|---|---|---|
| Alcohol (drinks per week) | Millwood *et al,* 2019 (14) | Mendelian randomization study |
| BMI | Carter *et al,* 2019 (7); Larsson *et al,* 2020 (15) | Mendelian randomization study; Mendelian randomization study |
| Low density lipoprotein cholesterol | Ference *et al,* 2018 (16) 2017;38(32):2459-72; Cheung *et al,* 2004 (17) | Mendelian randomization and clinical study; randomized controlled trial |
| Smoking (lifetime behaviour) | Larsson *et al,* 2020 (18) | Mendelian randomization study |
| Systolic blood pressure | Carter *et al,* 2019 (7); Ettehad *et al,* 2016 (19), | Mendelian randomization study; systematic review including randomized controlled trial |
| Type 2 diabetes | Ahmad *et al,* 2015 (20) | Mendelian randomization study |

**sTable 3: ICD codes for** coronary artery disease, stroke and type 2 **diabetes**

| Diagnosis | ICD9 | ICD10 |
|---|---|---|
| **Atrial Fibrillation** | 42731 | I48 |
| **Coronary heart disease** | 4100 - 4149 | I20-I25 |
| **Stroke** | 4300 - 4389 | I6, G45 |
| **Type 2 diabetes** | 4359 | G45 |

sTable 4: ICD codes for cardiovascular exclusions

| Cardiovascular event | ICD9 | ICD10 |
|---|---|---|
| **Myocardial infarction** | 4100-4109, 4120-4129 | I21, I22 |
| **Angina** | 4139 | I20 |
| **Transient ischaemic attack** | 4359 | G45 |
| **Peripheral arterial disease** | 4439 | I73.9 |
| **Stroke** | 4349 | I6, G45 |
| **Type 1 diabetes** | 2500- 25011, 25013, 2504-25041, 25043, 2505-25051, 25053, 2506-25061, 25063, 2507-25071, 25073, 2509-25091, 25093 | E10 |
| **Chronic kidney disease** | 5383, 5384, 5385 | N183, N184, N185 |
| **Familial hypercholesterolaemia** | 2720 | I78.0 |

sTable 5: Number of single nucleotide polymorphisms (SNPs) and variance explained ($R^2$) by polygenic scores for cardiovascular risk factors and outcomes

| | $P \leq 5x10^{-8}$ | | $P \leq 0.05$ | | $P \leq 0.5$ | |
|---|---|---|---|---|---|---|
| | $N_{SNPs}$ | $R^2$ | $N_{SNPs}$ | $R^2$ | $N_{SNPs}$ | $R^2$ |
| **Alcohol (drinks per week)** | 14 | 0.0840 | 72 962 | 0.0857 | 449 080 | 0.0857 |
| **Body mass index** | 127 | 0.0276 | 20 542 | 0.0646 | 139 582 | 0.0674 |
| **Low density lipoprotein cholesterol** | 398 | 0.0540 | 23 724 | 0.0144 | 13 337 | 0.0136 |
| **Systolic blood pressure (sample 1 GWAS)** | 126 | 0.1407 | 77 709 | 0.1579 | 373 402 | 0.1574 |
| **Systolic blood pressure (sample 2 GWAS)** | 112 | 0.1426 | 76 557 | 0.1633 | 372 715 | 0.1606 |
| **Smoking (sample 1 GWAS)** | 23 | 0.0097 | 67 741 | 0.0200 | 391 104 | 0.0206 |
| **Smoking (sample 2 GWAS)** | 21 | 0.0113 | 66 909 | 0.0222 | 390 557 | 0.0223 |
| **Atrial fibrillation** | 341 | 0.0061 | 60 738 | 0.0655 | 361 969 | 0.1064 |
| **Coronary heart disease** | 82 | 0.0654 | 49 098 | 0.0661 | 345 040 | 0.0656 |
| **Type 2 diabetes** | 18 | 0.0411 | 5 137 | 0.0366 | 134 673 | 0.0350 |
| **Stroke** | 11 | 0.0474 | 63 025 | 0.0480 | 373 240 | 0.0472 |

sTable 6: Association between per unit increase in educational attainment and observed phenotypic trait adjusted for age and sex

| **Trait** | **Mean difference in SD of phenotypic trait (95% CI)** |
|---|---|
| Alcohol (drinks per week) | 0.010 (0.009, 0.011) |
| BMI | -0.020 (-0.021, -0.020) |
| Low density lipoprotein cholesterol | $5.1x10^{-4}$ ($-2.0x10^{-4}$, $1.2x10^{-3}$) |
| Smoking (lifetime behaviour) | -0.028 (-0.028, -0.027) |
| Systolic blood pressure | -0.012 (-0.012, -0.011) |
| | **Risk difference of outcome (95% CI)** |
| Atrial fibrillation | $-5.1x10^{-4}$ ($-6.3x10^{-4}$, $-3.9x10^{-4}$) |
| Coronary artery disease | $-1.6x10^{-3}$ ($-1.7x10^{-3}$, $-1.5x10^{-3}$) |
| Diabetes (type 2) | $-1.7x10^{-3}$ ($-1.8x10^{-3}$, $-1.6x10^{-3}$) |
| Stroke | $-5.7x10^{-4}$ ($-6.6x10^{-4}$, $-4.7x10^{-4}$) |

sTable 7: Association between polygenic scores for susceptibility to continuous cardiovascular risk factors and phenotypic measure of each risk factor, stratified by educational attainment demonstrating effect modification

| Trait | Years of education | N | Additive scale | | Multiplicative scale | |
|---|---|---|---|---|---|---|
| | | | **Mean difference in SD of phenotypic trait (95% CI)** | **P value for effect modification** | **Mean difference in SD of log phenotypic trait (95% CI)** | **P value for effect modification** |
| Alcohol | All | 318 300 | 0.046 (0.042, 0.049) | | 0.049 (0.046, 0.053) | |
| | Education 7 | 51 509 | 0.037 (0.029, 0.046) | 0.384 | 0.040 (0.031, 0.049) | 0.976 |
| | Education 10 | 54 567 | 0.05 (0.042, 0.058) | | 0.058 (0.049, 0.066) | |
| | Education 13 | 17 267 | 0.045 (0.03, 0.059) | | 0.052 (0.037, 0.066) | |
| | Education 15 | 38 974 | 0.051 (0.041, 0.06) | | 0.056 (0.046, 0.065) | |
| | Education 19 | 51 095 | 0.05 (0.041, 0.059) | | 0.047 (0.039, 0.056) | |
| | Education 20 | 104 888 | 0.045 (0.039, 0.05) | | 0.048 (0.043, 0.053) | |
| | Effect modification coefficient | | $2.93x10^{-4}$ ($-3.66x10^{-4}$, $9.51x10^{-4}$) | | $9.96x10^{-4}$ ($-6.51x10^{-4}$, $6.71x10^{-4}$) | |
| BMI | All | 319 201 | 0.128 (0.124, 0.131) | | 0.128 (0.125, 0.131) | |
| | Education 7 | 51 773 | 0.129 (0.12, 0.138) | 0.036 | 0.126 (0.118, 0.135) | 0.330 |
| | Education 10 | 54 739 | 0.133 (0.125, 0.142) | | 0.132 (0.124, 0.14) | |
| | Education 13 | 17 319 | 0.131 (0.116, 0.146) | | 0.131 (0.116, 0.146) | |
| | Education 15 | 39 041 | 0.129 (0.119, 0.138) | | 0.129 (0.119, 0.138) | |
| | Education 19 | 51 309 | 0.136 (0.128, 0.145) | | 0.134 (0.126, 0.143) | |
| | Education 20 | 105 020 | 0.117 (0.111, 0.122) | | 0.121 (0.115, 0.126) | |
| | Effect modification coefficient | | $-7.20x10^{-4}$($-1.40x10^{-3}$, $-4.90x10^{-5}$) | | $-3.34x10^{-4}$ ($-1.01x10^{-3}$, $3.37x10^{-4}$) | |
| Low density lipoprotein cholesterol | All | 304 700 | 0.211 (0.208, 0.215) | | 0.209 (0.206, 0.212) | |
| | Education 7 | 49 435 | 0.186 (0.177, 0.195) | $1.12x10^{-4}$ | 0.18 (0.171, 0.189) | $1.63x10^{-6}$ |
| | Education 10 | 52 311 | 0.219 (0.211, 0.227) | | 0.216 (0.208, 0.224) | |
| | Education 13 | 16 521 | 0.224 (0.209, 0.238) | | 0.221 (0.207, 0.236) | |
| | Education 15 | 37 257 | 0.209 (0.199, 0.219) | | 0.206 (0.196, 0.215) | |
| | Education 19 | 48 942 | 0.206 (0.197, 0.214) | | 0.204 (0.196, 0.213) | |
| | Education 20 | 100 234 | 0.222 (0.216, 0.227) | | 0.221 (0.215, 0.227) | |
| | Effect modification coefficient | | $1.34x10^{-3}$ ($6.62x10^{-4}$, $2.02x10^{-3}$) | | $1.67x10^{-3}$ ($9.86x10^{-4}$, $2.35x10^{-3}$) | |
| Smoking (lifetime behaviour) | All | 301 684 | 0.039 (0.035, 0.042) | | 0.039 (0.035, 0.042) | |
| | Education 7 | 48 470 | 0.053 (0.042, 0.064) | 0.001 | 0.05 (0.039, 0.061) | 0.008 |
| | Education 10 | 52 292 | 0.035 (0.026, 0.044) | | 0.036 (0.027, 0.044) | |
| | Education 13 | 16 261 | 0.026 (0.012, 0.041) | | 0.026 (0.011, 0.041) | |
| | Education 15 | 37 149 | 0.046 (0.036, 0.055) | | 0.046 (0.036, 0.055) | |
| | Education 19 | 48 639 | 0.042 (0.032, 0.051) | | 0.042 (0.033, 0.052) | |
| | Education 20 | 98 873 | 0.029 (0.024, 0.034) | | 0.03 (0.025, 0.035) | |
| | Effect modification coefficient | | $-1.08x10^{-3}$ (-0.02, $-3.70x10^{-4}$) | | $-9.49x10^{-4}$ ($-1.64x10^{-3}$, - $2.53x10^{-4}$) | |
| Systolic blood pressure | All | 292 277 | 0.089 (0.086, 0.092) | | 0.070 (0.066, 0.074) | |
| | Education 7 | 46 726 | 0.080 (0.071, 0.089) | 0.104 | 0.058 (0.050, 0.067) | 0.076 |
| | Education 10 | 50 789 | 0.087 (0.079, 0.095) | | 0.069 (0.06, 0.079) | |
| | Education 13 | 15 772 | 0.094 (0.080, 0.108) | | 0.081 (0.064, 0.097) | |
| | Education 15 | 36 033 | 0.099 (0.089, 0.108) | | 0.078 (0.067, 0.088) | |
| | Education 19 | 47 177 | 0.088 (0.08, 0.097) | | 0.068 (0.058, 0.077) | |
| | Education 20 | 95 780 | 0.090 (0.084, 0.096) | | 0.073 (0.066, 0.08) | |
| | Effect modification coefficient | | $6.97x10^{-4}$ ($1.03x10^{-5}$, $1.38x10^{-3}$) | | $6.74x10^{-4}$ ($-7.03x10^{-5}$, $1.42x10^{-3}$) | |

sTable 8: Association between polygenic scores for susceptibility to cardiovascular risk factors and diseases and phenotypic measure of each risk factor or disease, stratified by educational attainment demonstrating effect modification

| Trait | Years of education | N | N cases | Additive scale | | Multiplicative scale | |
|---|---|---|---|---|---|---|---|
| | | | | **Risk difference (95% CI)** | **P value for effect modification** | **OR (95% CI)** | **P value for effect modification** |
| Atrial fibrillation | All | 316 912 | 9 560 | 0.0137 (0.0131, 0.0143) | | 1.59 (1.55, 1.62) | |
| | Education 7 | 51 246 | 2 438 | 0.0188 (0.017, 0.0206) | $9.03x10^{-08}$ | 1.51 (1.45, 1.57) | 0.008 |
| | Education 10 | 54 460 | 1 466 | 0.0123 (0.0109, 0.0136) | | 1.59 (1.51, 1.67) | |
| | Education 13 | 17 213 | 446 | 0.0126 (0.0102, 0.015) | | 1.66 (1.51, 1.82) | |
| | Education 15 | 38 694 | 1 232 | 0.0147 (0.0129, 0.0164) | | 1.60 (1.51, 1.69) | |
| | Education 19 | 50 942 | 1 396 | 0.0125 (0.0111, 0.0139) | | 1.58 (1.5, 1.67) | |
| | Education 20 | 104 357 | 2 582 | 0.0123 (0.0113, 0.0132) | | 1.65 (1.59, 1.71) | |
| | Effect modification coefficient | | | $-3.20x10^{-4}$ ($-4.30x10^{-4}$, $-2.00x10^{-4}$) | | 1.00 (1.00, 1.01) | |
| Coronary heart disease | All | 317 055 | 14 481 | 0.0085 (0.0077, 0.0092) | | 1.22 (1.2, 1.24) | |
| | Education 7 | 51 061 | 3 989 | 0.0115 (0.0091, 0.0138) | 0.103 | 1.18 (1.14, 1.21) | 0.001 |
| | Education 10 | 54 483 | 2 292 | 0.0079 (0.0062, 0.0095) | | 1.22 (1.17, 1.28) | |
| | Education 13 | 17 220 | 581 | 0.0053 (0.0027, 0.008) | | 1.18 (1.09, 1.28) | |
| | Education 15 | 38 740 | 1 733 | 0.0073 (0.0053, 0.0094) | | 1.19 (1.14, 1.25) | |
| | Education 19 | 50 912 | 2 477 | 0.0087 (0.0069, 0.0106) | | 1.22 (1.17, 1.27) | |
| | Education 20 | 104 639 | 3 409 | 0.0082 (0.0071, 0.0092) | | 1.30 (1.26, 1.35) | |
| | Effect modification coefficient | | | $-1.20x10^{-4}$ ($-2.60x10^{-4}$, $2.39x10^{-5}$) | | 1.00 (1.00, 1.01) | |
| Diabetes (Type 2) | All | 316 406 | 11 079 | 0.0081 (0.0074, 0.0087) | | 1.27 (1.24, 1.29) | |
| | Education 7 | 50 904 | 3 175 | 0.0131 (0.011, 0.0152) | $3.23x10^{-10}$ | 1.25 (1.2, 1.29) | 0.537 |
| | Education 10 | 54 261 | 1 809 | 0.0088 (0.0073, 0.0103) | | 1.31 (1.25, 1.37) | |
| | Education 13 | 17 190 | 512 | 0.0062 (0.0037, 0.0087) | | 1.23 (1.13, 1.35) | |
| | Education 15 | 38 683 | 1 336 | 0.0074 (0.0056, 0.0092) | | 1.24 (1.18, 1.31) | |
| | Education 19 | 50 814 | 1 914 | 0.0087 (0.007, 0.0103) | | 1.26 (1.21, 1.32) | |
| | Education 20 | 104 554 | 2 333 | 0.0057 (0.0048, 0.0066) | | 1.29 (1.24, 1.35) | |
| | Effect modification coefficient | | | $-4.00x10^{-4}$ ($-5.30x10^{-4}$, $-2.80x10^{-4}$) | | 1.00 (1.00, 1.00) | |
| Stroke | NONE | 320 120 | 5 929 | 0.0009 (0.0005, 0.0014) | | 1.05 (1.03, 1.08) | |
| | Education 7 | 52 012 | 1 620 | 0.002 (0.0005, 0.0035) | 0.036 | 1.07 (1.02, 1.12) | 0.292 |
| | Education 10 | 54 899 | 948 | 0.0007 (-0.0004, 0.0018) | | 1.04 (0.98, 1.11) | |
| | Education 13 | 17 355 | 311 | 0.0026 (0.0006, 0.0045) | | 1.16 (1.03, 1.29) | |
| | Education 15 | 39 144 | 731 | 0.0009 (-0.0005, 0.0022) | | 1.05 (0.97, 1.13) | |
| | Education 19 | 51 418 | 874 | 0.0016 (0.0005, 0.0027) | | 1.10 (1.03, 1.17) | |
| | Education 20 | 105 292 | 1 445 | $-4.83x10^{-5}$ (-0.0007, 0.0007) | | 1.00 (0.95, 1.05) | |
| | Effect modification coefficient | | | $-9.80x10^{-5}$ ($-1.90x10^{-4}$, $-6.40x10^{-6}$) | | 1.00 (0.99, 1.00) | |

sTable 9: Association between polygenic scores for susceptibility to continuous cardiovascular risk factors and phenotypic measure of each risk factor, stratified by educational attainment demonstrating effect modification using polygenic scores at a range of P value thresholds

| Exposure | Educational attainment | N | P≤0.05 | | | | P≤0.5 | | | |
|---|---|---|---|---|---|---|---|---|---|---|
| | | | Additive scale | | Multiplicative scale | | Additive scale | | Multiplicative scale | |
| | | | Mean difference in SD of phenotypic trait (95%CI) | P value for effect modification | Mean difference in SD of log phenotypic trait (95%CI) | P value for effect modification | Mean difference in SD of phenotypic trait (95%CI) | P value for effect modification | Mean difference in SD of log phenotypic trait (95%CI) | P value for effect modification |
| Alcohol | All years | 318,300 | 0.063 (0.06, 0.066) | | 0.064 (0.061, 0.068) | | 0.064 (0.061, 0.068) | | 0.065 (0.062, 0.069) | |
| | Education 7 | 51,509 | 0.056 (0.048, 0.065) | 0.694 | 0.062 (0.053, 0.071) | 0.108 | 0.058 (0.05, 0.067) | 0.669 | 0.065 (0.056, 0.074) | 0.04 |
| | Education 10 | 54,567 | 0.069 (0.061, 0.077) | | 0.07 (0.062, 0.079) | | 0.069 (0.061, 0.077) | | 0.071 (0.062, 0.079) | |
| | Education 13 | 17,267 | 0.075 (0.06, 0.091) | | 0.075 (0.06, 0.09) | | 0.072 (0.057, 0.088) | | 0.075 (0.06, 0.09) | |
| | Education 15 | 38,974 | 0.064 (0.054, 0.073) | | 0.065 (0.055, 0.075) | | 0.069 (0.06, 0.079) | | 0.069 (0.059, 0.079) | |
| | Education 19 | 51,095 | 0.056 (0.047, 0.065) | | 0.058 (0.049, 0.067) | | 0.061 (0.052, 0.07) | | 0.062 (0.053, 0.071) | |
| | Education 20 | 104,888 | 0.063 (0.057, 0.069) | | 0.06 (0.055, 0.066) | | 0.062 (0.056, 0.068) | | 0.059 (0.053, 0.065) | |
| | Effect modification coefficient | | -1.31x10$^{-4}$ (-7.85x10$^{-4}$, 5.23x10$^{4}$) | | -5.39x10-4 (-1.19x10-3, 1.18x10-4) | | -1.43x10$^{-4}$ (-7.95x10$^{-4}$, 5.10x10$^{-4}$) | | -6.68x10$^{-4}$ (-1.32x10$^{-3}$, -1.25x10$^{-5}$) | |
| Body mass index | All years | 319,201 | 0.229 (0.225, 0.232) | | -0.018 (-0.021, -0.014) | | 0.235 (0.231, 0.238) | | 0.237 (0.233, 0.24) | |
| | Education 7 | 51,773 | 0.225 (0.216, 0.233) | 0.005 | 0.220 (0.212, 0.229) | 0.453 | 0.228 (0.219, 0.237) | 0.215 | 0.224 (0.215, 0.232) | 0.396 |
| | Education 10 | 54,739 | 0.239 (0.231, 0.248) | | 0.239 (0.231, 0.247) | | 0.24 (0.232, 0.249) | | 0.24 (0.232, 0.249) | |
| | Education 13 | 17,319 | 0.235 (0.22, 0.25) | | 0.238 (0.223, 0.253) | | 0.245 (0.23, 0.26) | | 0.250 (0.235, 0.264) | |
| | Education 15 | 39,041 | 0.234 (0.224, 0.243) | | 0.235 (0.226, 0.245) | | 0.235 (0.225, 0.244) | | 0.236 (0.227, 0.246) | |
| | Education 19 | 51,309 | 0.235 (0.226, 0.243) | | 0.233 (0.225, 0.241) | | 0.238 (0.23, 0.247) | | 0.237 (0.228, 0.245) | |
| | Education 20 | 105,020 | 0.210 (0.204, 0.215) | | 0.217 (0.212, 0.223) | | 0.221 (0.216, 0.227) | | 0.229 (0.224, 0.235) | |
| | Effect modification coefficient | | -9.56x10$^{-4}$ (-1.62x10$^{-3}$, -2.96x10$^{-4}$) | | -2.52x10-4 (-9.10x10-4, 4.06x10-4) | | -4.17x10$^{-4}$ (-1.07x10$^{-3}$, 2.42x10$^{-4}$) | | 2.85x10$^{-4}$ (-3.72x10$^{-4}$, 9.42x10$^{-4}$) | |
| Low density lipoprotein cholesterol | All years | 304,700 | 0.071 (0.068, 0.075) | | 0.056 (0.052, 0.059) | | 0.065 (0.062, 0.069) | | 0.063 (0.06, 0.067) | |
| | Education 7 | 49,435 | 0.059 (0.05, 0.068) | 0.148 | 0.056 (0.047, 0.065) | 0.056 | 0.052 (0.043, 0.062) | 0.072 | 0.050 (0.04, 0.059) | 0.033 |
| | Education 10 | 52,311 | 0.08 (0.071, 0.088) | | 0.077 (0.068, 0.085) | | 0.073 (0.065, 0.082) | | 0.071 (0.062, 0.079) | |
| | Education 13 | 16,521 | 0.069 (0.054, 0.084) | | 0.066 (0.051, 0.081) | | 0.069 (0.055, 0.084) | | 0.065 (0.05, 0.08) | |
| | Education 15 | 37,257 | 0.069 (0.059, 0.08) | | 0.067 (0.057, 0.077) | | 0.057 (0.047, 0.067) | | 0.054 (0.044, 0.064) | |
| | Education 19 | 48,942 | 0.069 (0.06, 0.078) | | 0.068 (0.059, 0.077) | | 0.066 (0.058, 0.075) | | 0.064 (0.056, 0.073) | |
| | Education 20 | 100,234 | 0.075 (0.069, 0.081) | | 0.074 (0.068, 0.08) | | 0.069 (0.063, 0.075) | | 0.068 (0.062, 0.074) | |
| | Effect modification coefficient | | 5.12x10$^{-4}$ (-1.82x10$^{-4}$, 1.21x10$^{-3}$) | | 6.79x10-4 (-1.61x10-5, 1.37x10-3) | | 6.38x10$^{-4}$ (-5.80x10$^{-5}$, 1.33x10$^{-3}$) | | 7.59x10$^{-4}$ (6.21x10$^{-5}$, 1.46x10$^{-3}$) | |

| | | | | | | | | | | |
|---|---|---|---|---|---|---|---|---|---|---|
| Smoking (lifetime behaviour) | All years | 301,684 | 0.133 (0.128, 0.137) | | 0.135 (0.13, 0.14) | | 0.150 (0.145, 0.155) | | 0.152 (0.147, 0.157) | |
| | Education 7 | 48,470 | 0.184 (0.169, 0.198) | 2.16x10$^{-52}$ | 0.178 (0.165, 0.192) | 6.10x10$^{-40}$ | 0.197 (0.181, 0.213) | 8.85x10$^{-50}$ | 0.192 (0.177, 0.208) | 9.83x10$^{-38}$ |
| | Education 10 | 52,292 | 0.136 (0.124, 0.147) | | 0.137 (0.125, 0.149) | | 0.155 (0.142, 0.168) | | 0.157 (0.144, 0.169) | |
| | Education 13 | 16,261 | 0.114 (0.095, 0.133) | | 0.119 (0.1, 0.139) | | 0.128 (0.107, 0.149) | | 0.132 (0.111, 0.154) | |
| | Education 15 | 37,149 | 0.120 (0.108, 0.132) | | 0.122 (0.11, 0.135) | | 0.136 (0.122, 0.149) | | 0.139 (0.125, 0.152) | |
| | Education 19 | 48,639 | 0.141 (0.128, 0.153) | | 0.142 (0.129, 0.154) | | 0.165 (0.151, 0.178) | | 0.166 (0.152, 0.179) | |
| | Education 20 | 98,873 | 0.090 (0.084, 0.097) | | 0.097 (0.09, 0.103) | | 0.101 (0.094, 0.108) | | 0.109 (0.101, 0.116) | |
| | Effect modification coefficient | | -6.21x10$^{-3}$ (-7.01x10$^{-3}$, -5.41x10$^{-3}$) | | -5.39x10-3 (-6.19x10-3, -4.59x10-3) | | -6.38x10$^{-3}$ (-7.22x10$^{-3}$, 5.54x10$^{-3}$) | | -5.52x10$^{-3}$ (-6.37x10$^{-3}$, -4.68x10$^{-3}$) | |
| Systolic blood pressure | All years | 292,277 | 0.193 (0.189, 0.197) | | 0.155 (0.150, 0.160) | | 0.204 (0.2, 0.209) | | 0.163 (0.158, 0.168) | |
| | Education 7 | 46,726 | 0.181 (0.171, 0.192) | 0.160 | 0.139 (0.129, 0.15) | 0.127 | 0.189 (0.177, 0.2) | 0.124 | 0.143 (0.131, 0.154) | 0.191 |
| | Education 10 | 50,789 | 0.198 (0.188, 0.207) | | 0.165 (0.154, 0.176) | | 0.21 (0.2, 0.221) | | 0.177 (0.165, 0.189) | |
| | Education 13 | 15,772 | 0.205 (0.188, 0.222) | | 0.167 (0.147, 0.187) | | 0.225 (0.207, 0.243) | | 0.183 (0.161, 0.205) | |
| | Education 15 | 36,033 | 0.191 (0.179, 0.202) | | 0.145 (0.133, 0.158) | | 0.2 (0.188, 0.212) | | 0.153 (0.139, 0.167) | |
| | Education 19 | 47,177 | 0.182 (0.173, 0.192) | | 0.144 (0.133, 0.155) | | 0.195 (0.184, 0.205) | | 0.151 (0.139, 0.163) | |
| | Education 20 | 95,780 | 0.197 (0.19, 0.204) | | 0.163 (0.155, 0.172) | | 0.208 (0.2, 0.215) | | 0.169 (0.16, 0.178) | |
| | Effect modification coefficient | | 5.62x10$^{-4}$ (-2.22x10$^{-4}$, 1.35x10$^{-3}$) | | 6.90x10-4 (-1.97x10-4, 1.58x10-3) | | 6.73x10$^{-4}$ (-1.84x10$^{-4}$, 1.53x10$^{-3}$) | | 6.45x10$^{-4}$ (-3.24x10$^{-4}$, 1.61x10$^{-3}$) | |

sTable 10: Association between polygenic scores for susceptibility to cardiovascular risk factors and diseases and phenotypic measure of each risk factor or disease, stratified by educational attainment demonstrating effect modification using polygenic scores at a range of P value thresholds

| Exposure | Educational attainment | N | P≤0.05 | | | | P≤0.5 | | | |
|---|---|---|---|---|---|---|---|---|---|---|
| | | | Additive interaction | | Multiplicative interaction | | Additive interaction | | Multiplicative interaction | |
| | | | Risk difference of phenotypic trait (95%CI) | P value for effect modification | Odds ratio of phenotypic trait (95%CI) | P value for effect modification | Risk difference of phenotypic trait (95%CI) | P value for effect modification | Odds ratio of phenotypic trait (95%CI) | P value for effect modification |
| Atrial fibrillation | All years | 316,912 | 0.0454 (0.0448, 0.046) | | 4.67 (4.56, 4.79) | | 0.0586 (0.058, 0.0592) | | 5.88 (5.73, 6.03) | |
| | Education 7 | 51,246 | 0.0658 (0.0641, 0.0675) | | 4.44 (4.22, 4.67) | | 0.0827 (0.081, 0.0844) | | 5.51 (5.23, 5.8) | |
| | Education 10 | 54,460 | 0.0401 (0.0387, 0.0414) | | 4.5 (4.23, 4.79) | | 0.0519 (0.0506, 0.0533) | | 5.58 (5.23, 5.95) | |
| | Education 13 | 17,213 | 0.0392 (0.0369, 0.0415) | | 4.88 (4.34, 5.48) | | 0.052 (0.0497, 0.0544) | | 6.49 (5.74, 7.35) | |
| | Education 15 | 38,694 | 0.0474 (0.0457, 0.0491) | $4.52 \times 10^{-112}$ | 4.71 (4.4, 5.05) | 0.004 | 0.0607 (0.059, 0.0624) | $4.97 \times 10^{-167}$ | 5.77 (5.37, 6.2) | $2.87 \times 10^{-4}$ |
| | Education 19 | 50,942 | 0.0427 (0.0413, 0.0441) | | 4.92 (4.61, 5.26) | | 0.0557 (0.0543, 0.0571) | | 6.4 (5.97, 6.86) | |
| | Education 20 | 104,357 | 0.0391 (0.0382, 0.04) | | 4.83 (4.6, 5.06) | | 0.0507 (0.0498, 0.0517) | | 6.15 (5.85, 6.46) | |
| | Effect modification coefficient | | $-1.31 \times 10^{-3}$ ($-1.42 \times 10^{-3}$, $-1.19 \times 10^{-3}$) | | 1.01 (1.00, 1.01) | | $-1.57 \times 10^{-3}$ ($-1.69 \times 10^{-3}$, $-1.46 \times 10^{-3}$) | | 1.01 (1.00, 1.01) | |
| Coronary heart disease | All years | 317,055 | 0.0088 (0.008, 0.0095) | | 1.23 (1.21, 1.25) | | 0.0084 (0.0076, 0.0091) | | 1.22 (1.2, 1.24) | |
| | Education 7 | 51,061 | 0.013 (0.0107, 0.0153) | | 1.2 (1.16, 1.24) | | 0.0132 (0.0109, 0.0155) | | 1.21 (1.17, 1.25) | |
| | Education 10 | 54,483 | 0.0085 (0.0068, 0.0102) | | 1.24 (1.19, 1.3) | | 0.0067 (0.005, 0.0084) | | 1.19 (1.14, 1.24) | |
| | Education 13 | 17,220 | 0.0078 (0.0051, 0.0104) | | 1.28 (1.18, 1.4) | | 0.0063 (0.0036, 0.009) | | 1.23 (1.13, 1.34) | |
| | Education 15 | 38,740 | 0.0083 (0.0062, 0.0103) | $8.79 \times 10^{-06}$ | 1.22 (1.16, 1.28) | 0.372 | 0.0076 (0.0056, 0.0097) | $3.03 \times 10^{-4}$ | 1.2 (1.14, 1.26) | 0.140 |
| | Education 19 | 50,912 | 0.0085 (0.0066, 0.0103) | | 1.21 (1.16, 1.26) | | 0.0078 (0.0059, 0.0096) | | 1.19 (1.14, 1.24) | |
| | Education 20 | 104,639 | 0.0069 (0.0059, 0.008) | | 1.26 (1.21, 1.3) | | 0.0073 (0.0063, 0.0084) | | 1.27 (1.23, 1.31) | |
| | Effect modification coefficient | | $-3.22 \times 10^{-4}$ ($-4.64 \times 10^{-4}$, $1.80 \times 10^{-4}$) | | 1.00 (1.00, 1.00) | | $-2.61 \times 10^{-4}$ ($-4.03 \times 10^{-4}$, $1.19 \times 10^{-4}$) | | 1.00 (1.00, 1.01) | |
| Diabetes (Type 2) | All years | 316,406 | 0.0042 (0.0036, 0.0049) | | 1.14 (1.11, 1.16) | | 0.0016 (0.0009, 0.0022) | | 1.05 (1.03, 1.07) | |

| | | | | | | | | | | |
|---|---|---|---|---|---|---|---|---|---|---|
| | Education 7 | 50,904 | 0.0061 (0.004, 0.0082) | | 1.11 (1.07, 1.15) | | 0.0017 (-0.0004, 0.0037) | | 1.03 (0.99, 1.07) | |
| | Education 10 | 54,261 | 0.0046 (0.0031, 0.0061) | | 1.15 (1.1, 1.21) | | 0.0024 (0.0009, 0.0039) | | 1.08 (1.03, 1.13) | |
| | Education 13 | 17,190 | 0.0028 (0.0002, 0.0053) | | 1.11 (1.01, 1.21) | | 0.0021 (-0.0004, 0.0047) | | 1.07 (0.98, 1.17) | |
| | Education 15 | 38,683 | 0.004 (0.0021, 0.0058) | 0.011 | 1.13 (1.07, 1.19) | 0.273 | 0.0009 (-0.0009, 0.0027) | 0.317 | 1.03 (0.97, 1.09) | 0.705 |
| | Education 19 | 50,814 | 0.0059 (0.0042, 0.0075) | | 1.18 (1.13, 1.24) | | 0.0018 (0.0002, 0.0035) | | 1.05 (1.01, 1.1) | |
| | Education 20 | 104,554 | 0.0027 (0.0019, 0.0036) | | 1.14 (1.09, 1.18) | | 0.0011 (0.0002, 0.002) | | 1.05 (1.01, 1.1) | |
| | Effect modification coefficient | | $-1.64\times10^{-4}$ ($-2.90\times10^{-4}$, $-3.81\times10^{-5}$) | | 1.00 (1.00, 1.01) | | $-6.41\times10^{-5}$ ($-1.90\times10^{-4}$, $6.15\times10^{-5}$) | | 1.00 (1.00, 1.00) | |
| Stroke | All years | 320,120 | 0.0016 (0.0012, 0.0021) | | 1.1 (1.07, 1.13) | | 0.0003 (-0.0001, 0.0008) | | 1.02 (0.99, 1.05) | |
| | Education 7 | 52,012 | 0.0033 (0.0018, 0.0048) | | 1.12 (1.06, 1.17) | | 0.0007 (-0.0008, 0.0022) | | 1.02 (0.97, 1.08) | |
| | Education 10 | 54,899 | 0.001 (-0.0001, 0.0021) | | 1.06 (0.99, 1.13) | | 0.0005 (-0.0006, 0.0016) | | 1.03 (0.97, 1.1) | |
| | Education 13 | 17,355 | 0.0005 (-0.0015, 0.0025) | | 1.03 (0.92, 1.16) | | 0.0002 (-0.0018, 0.0022) | | 1.01 (0.9, 1.14) | |
| | Education 15 | 39,144 | 0.0024 (0.0011, 0.0038) | 0.015 | 1.15 (1.06, 1.24) | 0.538 | 0.0004 (-0.0009, 0.0018) | 0.378 | 1.02 (0.95, 1.1) | 0.666 |
| | Education 19 | 51,418 | 0.0011 (0, 0.0023) | | 1.07 (1, 1.15) | | -0.0006 (-0.0017, 0.0005) | | 0.97 (0.9, 1.03) | |
| | Education 20 | 105,292 | 0.0013 (0.0006, 0.002) | | 1.1 (1.04, 1.16) | | 0.0005 (-0.0002, 0.0012) | | 1.04 (0.99, 1.1) | |
| | Effect modification coefficient | | $-1.31\times10^{-4}$ ($-2.05\times10^{-4}$, $-2.17\times10^{-5}$) | | 1.00 (0.99, 1.00) | | $-4.13\times10^{-5}$ ($-1.33\times10^{-4}$, $5.05\times10^{-5}$) | | 1.00 (0.99, 1.00) | |
